# Levels and determinants of child wasting relapse: a prospective cohort study from Somalia

**DOI:** 10.1101/2025.04.18.25326026

**Authors:** Kemish Kenneth Alier, Shelley Walton, Samantha Grounds, Sydney Garretson, Said Aden Mohamoud, Mohamud Ali Nur, Sadiq Abdiqadir, Mohamed Billow Mahat, Michael Ocircan P’Rajom, Meftuh Omer Ismail, Abdullahi Farah, Qundeel Khattak, Lilly Schofield, Marina Tripaldi, Fabrizio Loddo, Pierluigi Sinibaldi, Farhan Mohamed, Abdifatah Ahmed Mohamed, Adam Abdulkadir Mohamed, Nadia Akseer

## Abstract

**Background:** Understanding the rates and determinants of severe acute malnutrition (SAM) relapse is crucial for stakeholders in Somalia, where evidence is limited. This study aimed to assess SAM relapse rates and associated risk factors among children discharged from outpatient therapeutic programs (OTP) in the Bay and Hiran regions of Somalia.

**Methods:** We conducted a prospective cohort study of 160 children aged 7-53 months discharged as recovered from OTP SAM treatment between August and September 2023. Children were followed monthly for 6 timepoints post-discharge. Anthropometric measurements, morbidity data, and household information were collected. Survival analysis was used to calculate cumulative incidence of SAM relapse, defined by weight-for-height z-score (WHZ) <-3 SD or mid-upper arm circumference (MUAC) <11.5cm or oedema. Cox proportional hazard models identified factors associated with relapse.

**Results:** Cumulative incidence of SAM relapse at T1=5.2% (CI: 2.5-10.6%)), T2=14.3% (9.4-21.5%) and T6 was 26.0% (CI: 19.3-34.5%) by WHZ and 13.2% (CI: 8.8-19.5%) by MUAC. The relapse rate for combined SAM and MAM by WHZ at T1=26.9% (CI: 19.5-36.3%), T2=36.2% (CI: 28.0-46.1%) and T6=50.1% (CI: 41.0-60.0%). WHZ-based relapse was higher in rural areas (31.4% vs 22.7% urban, p=0.285) and among children with WHZ <-3SD at admission (37.4% vs 21.2%, p=0.029). MUAC-based relapse was higher in urban areas (20.8% vs 4.1% rural, p=0.002), among younger children (19.7% vs 5.5% >2 years, p=0.009), and IDPs (21.8% vs 5.8% non-IDPs, p=0.003). Factors significantly associated with increased relapse risk included WHZ <-3 SD at admission (adjusted HR: 2.22, CI: 1.04-4.72) and longer OTP stay (adjusted HR: 1.02 per day, CI: 1.00-1.04). Participation in a cash assistance program was protective (adjusted HR: 0.44, CI: 0.22-0.90).

**Conclusions:** SAM relapse rates in Somalia are considerable, and varies by indicators, regions, and demographics. cash assistance shows promise for improving outcomes.

**Registration:** The cluster-RCT associated with this cohort study is registered at ClinicalTrials.gov, ID: NCT06642012.

## BACKGROUND

Acute malnutrition, also known as wasting, is a major global health concern, impacting an estimated 45 million children under 5 (CU5) worldwide [1]. While progress has been made against other indicators of malnutrition, such as stunting, the prevalence of wasting has not seen a similar decline and is a significant contributor to CU5 morbidity and mortality [2]. In high-burden settings, children are screened for wasting by community health workers, family members and health workers and referred to Integrated Management of Acute Malnutrition (IMAM) programs for treatment [3]. Once a child achieves recovery criteria (WHZ ≥ −2 and/or MUAC ≥ 125 mm and no oedema for at least 2 weeks), they are discharged from the program [4]. While most children treated for SAM successfully recover, they can experience higher nutritional vulnerability post-treatment than their never-wasted counterparts. In a matched cohort study of CU5 in Ethiopia, post-SAM children experienced a greater burden of illness and relapse than healthy peers in the same village [5]. Recent systematic reviews show that child wasting relapse rates vary widely from 0%-37% globally, with follow-up times ranging from 1 week to 18 months, and relapse risk highest within the first 3-6 months [6]. Post-discharge mortality is also highest among post-SAM children.[7]

Somalia, an impoverished country of 18 million people, struggles with conflict, environmental disasters, extreme food insecurity and high rates of malnutrition [8]. Understanding relapse rates and determinants is a top priority, but evidence is limited. A recent multi-country study found that 5% of SAM-recovered children in Somalia either relapsed according to MUAC-only criteria or died within six months. When MUAC and/or WHZ criteria were applied, relapse increased to 23%[9]. However, this study was limited to one nutrition site in urban Mogadishu, the capital of Somalia. [10] [11] While frameworks exist for understanding drivers of wasting relapse, and some empirical work has been done in settings such as Nepal, Niger, Ethiopia, Mali, and Nigeria, evidence from Somalia is sparse [11], [12–16]. Since risk and protective factors vary notably by context, understanding factors affecting relapse in Somalia is crucial for effective policy and programming. This study seeks to address this evidence gap by estimating wasting relapse rates, particularly SAM relapse rate and determinants among children under 5 years discharged from outpatient therapeutic feeding programs in two high-burden settings of Somalia, Bay and Hiran. Evidence from this study could inform funding, policy and programming efforts for identifying and treating children at high risk of relapse in Somalia and similar humanitarian settings.

## METHODS

### Study setting

During our study, Somalia ranked first on the Fragile States Index [17] with more than half of the 18 million population living in poverty [18]. Food insecurity is highly prevalent with up to 1 in 5 Somalis classified by Integrated Food Security Phase Classification (IPC) as facing severe hunger in 2024 and over 1.7 million under five children were acutely malnourished [19]. This study was conducted in the Bay and Hiran regions of southwestern Somalia. These two regions were selected due to the high prevalence of acute malnutrition and food insecurity. Additionally, a Bureau for Humanitarian Assistance (BHA) program (supported by USAID), which focused on providing life-saving cash for nutrition to communities, was being implemented in these regions.

The Bay region has a semi-arid climate and is prone to recurrent droughts and floods, which negatively impact agricultural production and food security. The region has a predominantly agro-pastoral economy, with many households relying on livestock rearing and crop cultivation as their primary livelihood activities [20]. The Hiran region experiences a semi-arid climate and is also vulnerable to climatic shocks, such as droughts and floods. The region’s economy is primarily based on pastoralism, with many households heavily dependent on livestock for their livelihoods [21]. Both the Bay and Hiran regions have been affected by years of conflict, insecurity, food crises, and environmental degradation, which have contributed to high levels of acute malnutrition among children under five years of age [22] and as represented in Figure 1 of acute malnutrition between October 2023-February 2024 [19].

**Figure 1:**
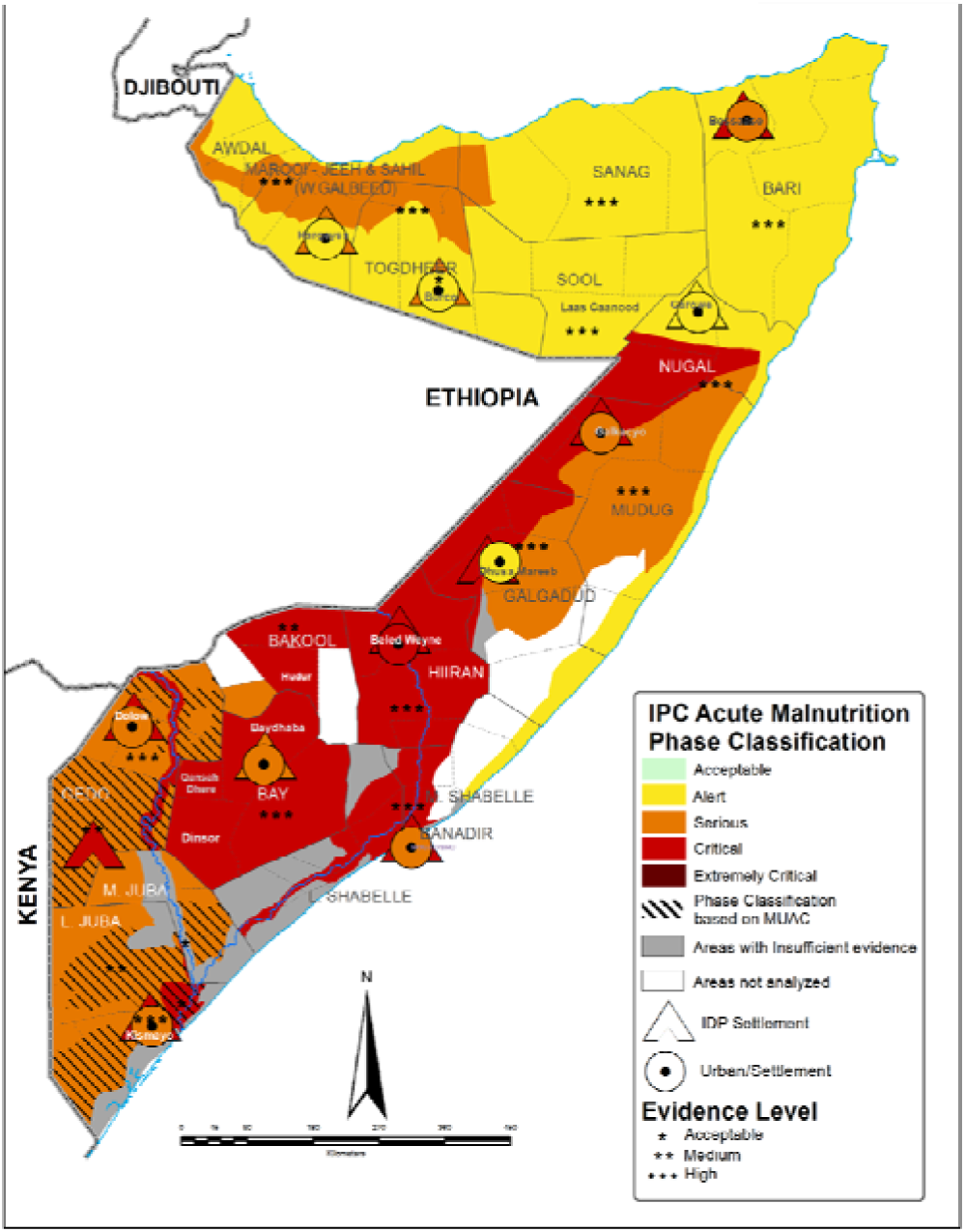
Map of Somalia showing burden of acute wasting by region (Oct 2023-Feb 2024). Source: IPC report[19]

### Study Design

This was a prospective cohort study for SAM children discharged from OTP and followed up for relapse. The study was conducted in Somalia between August 2023 to April 2024 in health facilities supported by Save the Children and follows the Somali guidelines for integrated management and treatment of acute malnutrition [23]. The OTP details are summarized in **Panel 1**.

**Panel 1:** Study Context and Outpatient Therapeutic Feeding Program (OTP) details.

| Category |  | Specification |
| --- | --- | --- |
| Context | Locations | Bay (7 OTP sites)<br>Hiran (14 OTP sites) |
|  | Population/Livelihood | IDPs, resident pastoral and agro-pastoral population |
|  | Urban, Rural Status | A mix of rural and urban locations |
|  | Implementer of health services | Ministry of Health and Save the Children |
| OTP Program Details | OTP Admission criteria | MUAC <11.5cm and/or WHZ < -3 SD and/or oedema |
|  | OTP products used | Ready-to-Use Therapeutic Food (RUTF) |
|  | OTP Discharge/transfer criteria | MUAC ≥ 11.5cm and/or WHZ ≥ -1.5 SD and no oedema<br>(if both MUAC and WHZ below threshold at admission, discharge is based on either one or both, whichever meets discharge criteria first) |
|  | Transfer from OTP to Targeted Supplementary Feeding Programme (TSFP) | Yes, but with modification in some cases. Treat SAM children until children transition from the SAM to MAM phase of treatment, at which point they are referred to TSFP for the remainder of treatment; if no TSFP available, will stay in OTP until fully recovered from wasting |
|  | OTP follow-up frequency | Weekly follow-up consultation visits at point of care |
| Post-Discharge | Definition of relapse |  |
|  | SAM relapse | Defined using one indicator: Either WHZ < -3 SD or MUAC <11.5cm |
|  | Both SAM+MAM relapse | Defined using one indicator: Either WHZ < -2SD or MUAC < 12.5 cm |

We employed a convenience sampling approach enrolling children who had been treated and discharged from an OTP program in the study regions. As this was a descriptive study, we aimed to reach a maximum sample size from all eligible children, as resources and time allowed.

We extracted the admission and discharge demographic and clinical data from registers of 21 health facilities (15 Hiran 7 Bay) for children discharged from OTP between August 24^th^ and September 19^th^, 2023, and screened them for enrollment **(Figure 2)**. Inclusion criteria included children having no medical complications, being aged 7-53 months at OTP exit, and being discharged with a recovered outcome based on Somalia IMAM guidelines (MUAC≥ 11.5cm and/or WHZ≥ −3 SD) and no oedema [23]. We excluded children with presence of a chronic or congenital disease (not including HIV infection or TB) or disability that affects growth, ability to consume food, or anthropometric measurements (e.g. cerebral palsy), or if the initial OTP admission was preceded by enrollment in an inpatient, hospital, or stabilization center where the child was being treated for SAM with medical complications similar to previous studies [10],[24].

**Figure 2:**
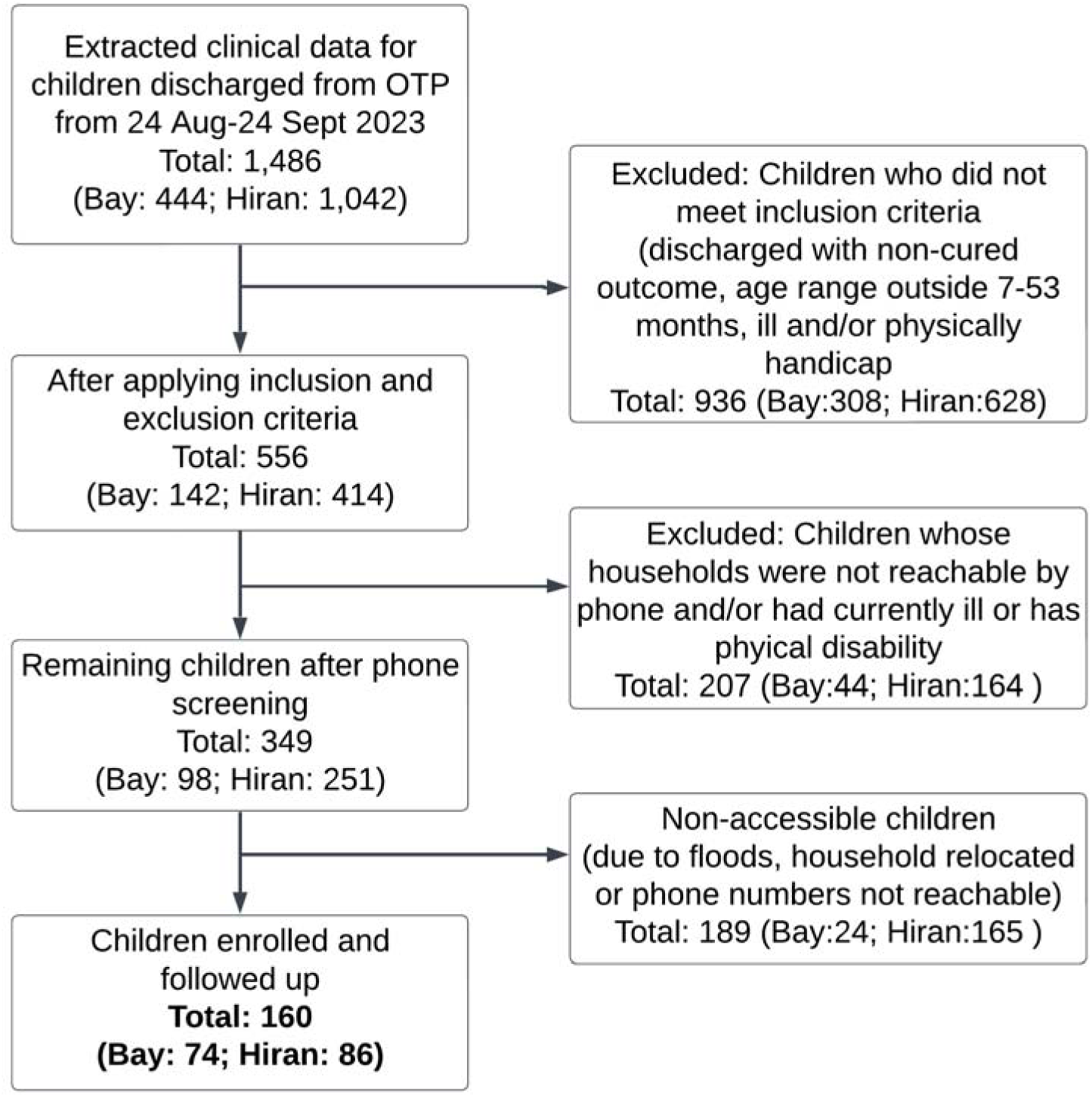
Study recruitment and follow-up flow chart

Subsequently, we phoned caregivers of the eligible children to determine additional inclusion information as follows: 1) presence of the household in the two study regions and if they intend on staying in the same location for the next 6 months; 2) to ascertain if the child has any chronic medical condition not captured in the medical register, and 3) if they were willing to participate in the study. The final study enrolled a total of 160 children (Bay=74, Hiran=86) who met all inclusion criteria and whose families provided consent to participate. Consent was confirmed at each follow up point.

### Data collection

Enrolled post-SAM children were followed up monthly at home for six follow-ups post-discharge to assess for SAM as well as linear growth, morbidity, and mortality outcomes. We aimed to follow up children soon after discharge (baseline). The average time for first follow-up post discharge was 60±15 days (hereafter Time 1(T1)). The second to the sixth follow-up were 30 days (one month) apart on average (hereafter Time 2, Time 3, Time 4, Time 5 and Time 6 (T2,T3,T4,T5, and T6)). The first follow-up was conducted from 18-26 November 2023 and the final follow-up was conducted from 15-20 April 2024 corresponding to the second raining season (deyr) and dry season (Jilaal). We use standard anthropometric measurement methods [25].

At the first follow-up, we collected detailed individual, household, and community-level information: 1) Individual: anthropometry, age, gender, illness in past two weeks, infant and young child feeding (IYCF), vaccine status, Minimum dietary diversity for children (MDD-C), wasting treatment program experience; 2) Household: maternal weight, household size/composition, decision making, assets, food consumption score, hygiene practices, mother education, status of household head, type of water source, household food security indictors (food consumption score, household hunger scale and reduced coping strategies index); and 3) Community: internally displaced person (IDP) status, recent shocks experience (floods) and whether household was receiving cash transfers from the BHA-supported project. The questionnaire is in **Table S1.** At subsequent follow-ups, only child anthropometry and morbidity data were collected.

### Study outcomes

The primary outcome of interest was cumulative incidence of SAM defined by WHZ or MUAC or oedema. We defined SAM relapse as children whose WHZ score falls below −3 SD during follow-up or MUAC measurement below 11.5cm and no oedema. Our secondary outcome was cumulative incidence of moderate acute malnutrition (MAM) for a sub-set of post-SAM children. We defined MAM relapse as any child whose WHZ score falls below −2 SD. We didn’t use MUAC as there were no children with MUAC >12.5cm at enrollment.

We used the zscore06 Stata module based on the 2006 WHO child growth standards to calculate anthropometric indices (WHZ, WAZ, HAZ).[26,27] Extreme values were flagged and excluded according to WHO recommendations: WHZ < −5 or > 5, WAZ < −6 or > 5, and HAZ < −6 or > 6. We constructed three analysis cohorts based on the MUAC and WHZ score at OTP exit: 1) cohort for WHZ ≥-3 SD at exit and followed up for relapse to WHZ <-3 SD (SAM), 2) cohort for MUAC ≥11.5cm at exit and followed up for relapse to MUAC < 11.5cm (SAM), and 3) cohort for WHZ ≥-2 SD at exit and followed up for relapse to WHZ <-2 SD (Both SAM+MAM). We created these cohorts because the heath facilities discharge criteria varied. While some use only MUAC, others used WHZ for discharging children from OTP. The criteria for discharge didn’t always align with criteria used for admission. The cohorts allowed us to examine the different exit scenarios.

### Statistical analysis

Descriptive statistics were calculated for baseline characteristics, including demographics, anthropometry, and household factors. Continuous variables were summarized as means and standard deviations, and categorical variables were presented as frequencies and percentages. We used t-tests for continuous variables and Chi-square tests for categorical variables to assess differences between regions. For each cohort, we conducted time-to-event analyses using Kaplan-Meier survival curves to estimate the cumulative incidence of relapse at 6 different timepoints (T1–T6) post-OTP exit. The log-rank test was used to compare survival curves across regions, age groups, urban/rural, and severity of wasting at admission. We calculated incidence rates of relapse with 95% confidence intervals.

To identify factors associated with relapse, we performed bivariate and multivariable Cox proportional hazards regression analyses. Variables with p<0.2 in bivariate analyses were included in the initial multivariate model. We used a backward stepwise approach to build the final model, retaining variables with p < 0.05. The proportional hazards assumption was tested using Schoenfeld residuals. We reported unadjusted and adjusted hazard ratios (HR) with 95% confidence intervals. The final model was adjusted for child age, sex, livelihood zone, and time to relapse. To account for potential clustering effects at the OTP site level, we used robust standard errors in our regression models. We conducted sensitivity analyses to assess the impact of different definitions of relapse using combinations of MUAC and WHZ and to evaluate the effect of children we were not able to reach after extraction of clinic data. All statistical tests were two-sided, and p-values < 0.05 were considered statistically significant. We followed STROBE guidelines for reporting observational studies.[28] Data were analyzed using Stata version 18 (StataCorp, College Station, TX, USA).

## RESULTS

A total of 160 post-SAM children were successfully recruited and followed for the duration of the study. There was no dropout during the follow-up period.

### Child and household characteristics

Post-SAM children were predominantly female and aged 2 years and below **(Table 1).**

**Table 1:** Characteristics of Children and Household at Baseline.

| Variable | Overall | By Region |  |  |
| --- | --- | --- | --- | --- |
|  |  | Bay | Hiran | p-value |
| Sample:(N/%) | 160 | 74 (46.2%) | 86 (53.8%) |  |
| Sex (n/%) |  |  |  |  |
| Male | 72 (45.0%) | 37 (50.0%) | 35 (40.7%) | 0.238 |
| Female | 88 (55.0%) | 37 (50.0%) | 51 (59.3%) |  |
| Age, months (n/%) |  |  |  |  |
| < 2 yrs | 82 (51.2%) | 56 (75.7%) | 26 (30.2%) | <0.001 |
| ≥ 2 yrs | 78 (48.8%) | 18 (24.3%) | 60 (69.8%) |  |
| **Household Characteristics** |  |  |  |  |
| Rural/Urban (n/%) |  |  |  |  |
| Rural | 73 (45.6%) | 0 (0.0%) | 73 (84.9%) | <0.001 |
| Urban | 87 (54.4%) | 74 (100.0%) | 13 (15.1%) |  |
| Livelihood zone (n/%) |  |  |  |  |
| Pastoral | 45 (28.1%) | 0 (0.0%) | 45 (52.3%) | <0.001 |
| Agropastoral | 41 (25.6%) | 0 (0.0%) | 41 (47.7%) |  |
| IDP | 74 (46.2%) | 74 (100.0%) | 0 (0.0%) |  |
| Save the Children BHA Program* (n/%) |  |  |  |  |
| Not in BHA Program | 56 (35.0%) | 33 (44.6%) | 23 (26.7%) | 0.018 |
| In BHA program | 104 (65.0%) | 41 (55.4%) | 63 (73.3%) |  |
| Shocks (Affected by floods during study) (n/%) |  |  |  |  |
| Not flood affected | 38 (30.4%) | 12 (16.4%) | 26 (50.0%) | <0.001 |
| Flood affected | 87 (69.6%) | 61 (83.6%) | 26 (50.0%) |  |
| **Admission Profile** |  |  |  |  |
| WHZ Score (mean/sd) | -2.5 (1.3) | -2.1 (1.1) | -2.8 (1.4) | <0.001 |
| WHZ ≥ -3 to < -2 (n/%) | 44 (28.0%) | 23 (31.1%) | 21 (25.3%) | |
| WHZ < -3 SD (n/%) | 63 (40.1%) | 15 (20.3%) | 48 (57.8%) |  |
| MUAC, cm (mean/sd) | 11.4 (0.7) | 11.1 (0.7) | 11.6 (0.6) | <0.001 |
| Wasting, MUAC (n/%) |  |  |  |  |
| ≥ 11.5 to < 12.5 | 30 (18.8%) | 3 (4.1%) | 27 (31.4%) | |
| < 11.5 | 121 (75.6%) | 71 (95.9%) | 50 (58.1%) |  |
| Oedema At Admission (n/%) |  |  |  |  |
| Yes | 6 (3.8%) | 0 (0.0%) | 6 (7.0%) | 0.021 |
| **Discharge/Enrolment Profile** |  |  |  |  |
| Length of stay, days (mean/sd) | 42.4 (18.3) | 34.2 (10.9) | 49.5 (20.4) | <0.001 |
| WHZ Score (mean/sd) | -1.4 (1.4) | -1.0 (1.1) | -1.7 (1.6) | <0.001 |
| MUAC, cm (mean/sd) | 12.0 (0.6) | 11.9 (0.2) | 12.1 (0.7) | 0.002 |
| Underweight, WAZ (n/%) |  |  |  |  |
| WAZ ≥ -3 to < -2 | 57 (35.6%) | 35 (47.3%) | 22 (25.6%) | 0.001 |
| WAZ < -3 SD | 31 (19.4%) | 17 (23.0%) | 14 (16.3%) |  |
| Stunting, HAZ (n/%) |  |  |  |  |
| HAZ ≥ -2 SD | 72 (45.9%) | 17 (23.0%) | 55 (66.3%) | <0.001 |
| HAZ < -2 SD | 85 (54.1%) | 57 (77.0%) | 28 (33.7%) |  |
| **IMAM program quality** |  |  |  |  |
| Child was not transferred from OTP to TSFP | 79 (49.4%) | 27 (36.5%) | 52 (60.5%) | 0.002 |
| Child was transferred from OTP to TSFP | 81 (50.6%) | 47 (63.5%) | 34 (39.5%) |  |

\* There was an ongoing BHA livelihood program implemented by Save the Children in the two regions for vulnerable households. The BHA interventions included multi-purpose cash transfers and SBCC interventions.

Notably, Bay region had a significantly higher proportion of younger children (81.1%) compared to Hiran (31.4%, p<0.001). Household characteristics differed between regions. Bay was exclusively urban (100%) and comprised entirely of IDPs, while Hiran was predominantly rural (84.9%) with a mix of pastoral (52.3%) and agropastoral (47.7%) livelihoods (p<0.001). More households were affected by the floods shock in Bay (83.6%) than Hiran (50.0%, p<0.001).

At admission, the overall mean WHZ was −2.5±1.3 SD, with Hiran showing more severe wasting (WHZ<-3: 57.8%) than Bay (20.3%, p<0.001). Mean MUAC at admission was 11.4±0.7 cm, and Bay (11.1±0.7 cm) had a lower mean MUAC than Hiran (11.6±0.6 cm, p<0.001). The overall length of stay (LOS) in OTP SAM treatment was 42.4±18.3 days, with children staying longer in Hiran (49.5±20.4 days) than Bay (34.2±10.9 days, p<0.001). Discharge WHZ was −1.4±1.4 SD overall, with Bay (−1.0±1.1 SD) having greater mean WHZ than Hiran (−1.7±1.6 SD, p<0.001). Mean MUAC at discharge was 12.0±0.6 cm, with slight regional variation (Bay: 11.9±0.2 cm, Hiran: 12.1±0.7 cm, p=0.002). Over half of the children were stunted (HAZ < −2 SD) at discharge. Almost half of the children (49.3%) were not transferred to TSFP treatment after exiting OTP.

### Relapse incidence

**Table 2** shows the relapse incidence by the analysis cohorts. For SAM relapse using WHZ, the highest incidence of relapse was in the first 3-4 months with 5.2%, 9.1%, 6.1% relapsing to SAM at T1, T2 and T3 respectively. From T4 to T6, relapse incidence rate was lower at 1.5%, 0.8% and 3.3% respectively. The cumulative incidence rate at T6 was 26.0% (95% CI 19.3-34.5%). Rural children had a higher relapse rate (31.4%, 95% CI 20.5-46.0%) compared to urban children at T6 (22.7%, 95% CI 14.9-33.7%), however this difference was not statistically significant (p=0.285). Hiran region had a higher relapse rate (30.2%) than Bay region (21.8%) at T6, but this difference was also not statistically significant (p=0.309). As shown in **Figure 3(a)**, older children (≥2 years) showed a slightly higher relapse rate (29.5%) than younger children (23.3%), but not statistically significant (p=0.488). Notably, in **Figure 3(b)**, children who had WHZ <-3 SD at admission had significantly higher relapse rates (37.4%) than children admission WHZ ≥ −3 SD (21.2%, p=0.029). Children who were not transferred from OTP to TSFP had a higher relapse rate at T6 (30.9% vs 21.7% for those referred to TSFP (p=0.237).

**Figure 3(a):**
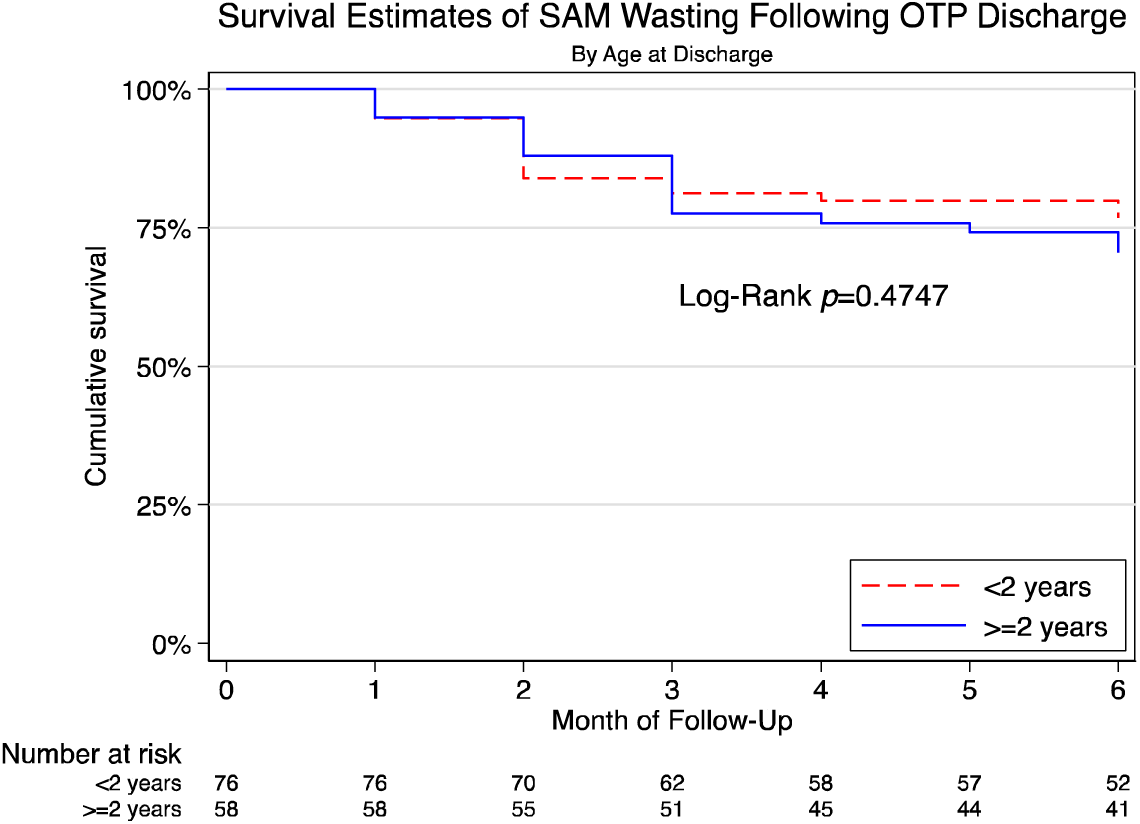
Survival Estimates of SAM wasting following OTP discharge (By age)

**Figure 3(b):**
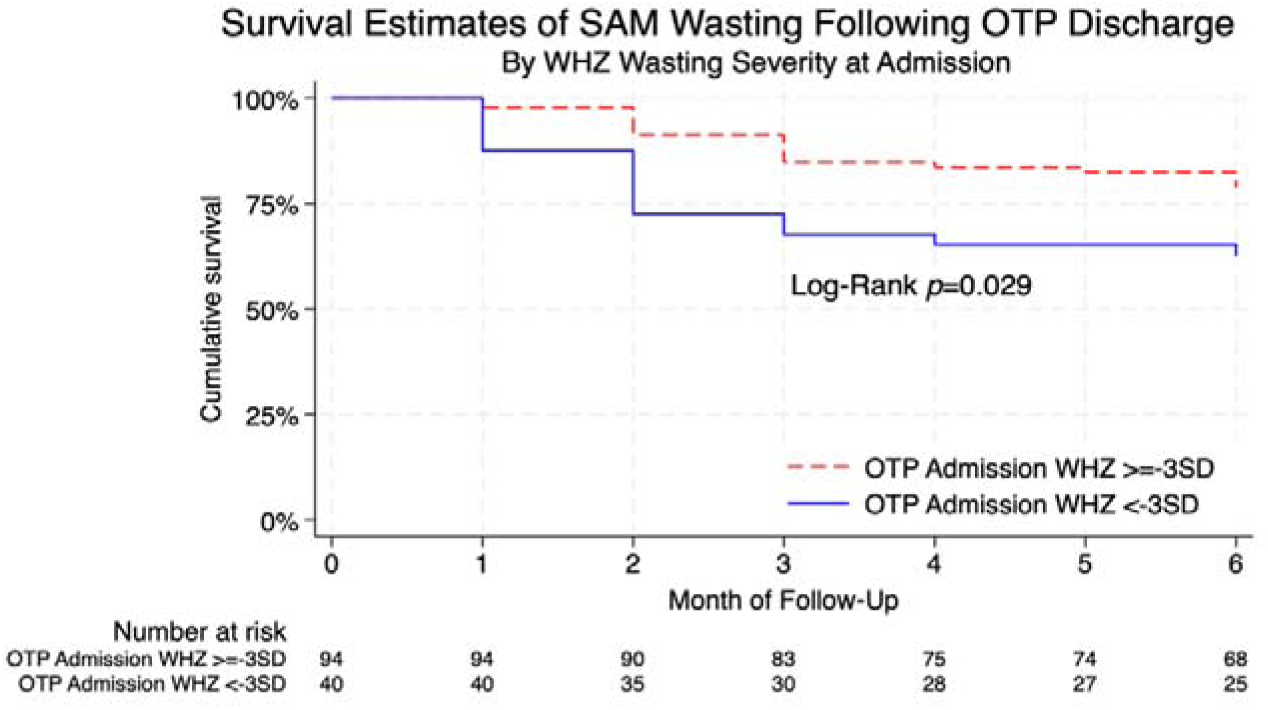
Survival Estimates of SAM wasting following OTP discharge (By wasting severity at admission)

**Figure 3(c):**
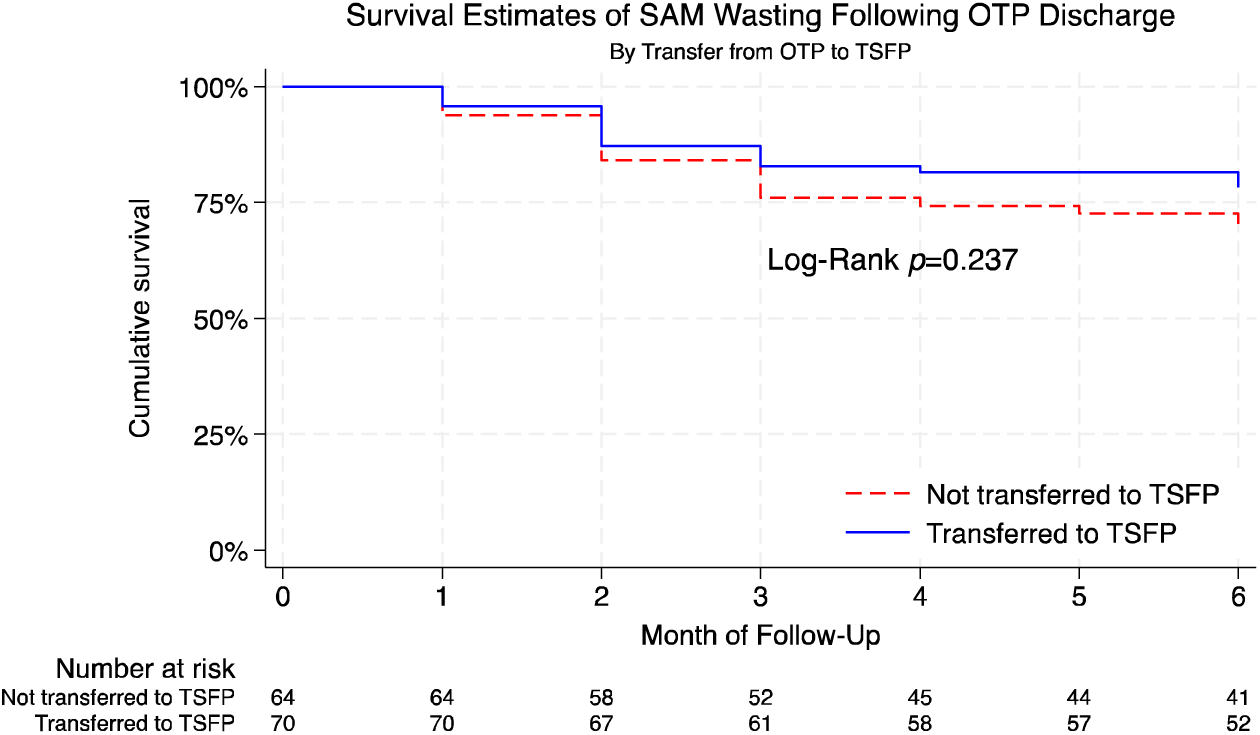
Survival Estimates of SAM wasting following OTP discharge (By whether the child was transferred from OTP to TSFP)

**Table 2:** Incidence of Wasting for SAM (By WHZ), SAM (By MUAC) and MAM (By WHZ) at each follow-up point.

|  | Overall |  | Region |  |  |  |  | Rural Urban |  |  |  |  |
| --- | --- | --- | --- | --- | --- | --- | --- | --- | --- | --- | --- | --- |
| Follow-up Time | Incidence | Cumulative Incidence (95% CI) | Incidence | Cumulative Incidence (95% CI) | Incidence | Cumulative Incidence (95% CI) | p-value | Incidence | Cumulative Incidence (95% CI) | Incidence | Cumulative Incidence (95% CI) | p-value |
| **SAM by WHZ** |  |  |  |  |  |  |  |  |  |  |  |  |
|  | (n=134) |  | Bay (IDP population) n=71 |  | Hiran (Non-IDP population) n=63 |  |  | Rural (n=51) |  | Urban (n=83) |  |  |
| T1 | 5.2% | 5.2% (2.5 - 10.6) | 4.2% | 4.2% (1.4 - 12.5) | 6.3% | 6.3% (2.4 - 16.0) |  | 5.9% | 5.9% (1.9 - 17.1) | 4.8% | 4.8% (1.8 - 12.3) |  |
| T2 | 9.1% | 14.3% (9.4 - 21.5) | 10.2% | 14.4% (8.0 - 25.1) | 8.0% | 14.3% (7.7 - 25.7) |  | 7.8% | 13.7% (6.8 - 26.7) | 9.9% | 14.7% (8.6 - 24.5) |  |
| T3 | 6.1% | 20.4% (14.5 - 28.3) | 4.3% | 18.7% (11.3 - 30.1) | 7.9% | 22.2% (13.8 - 34.6) |  | 9.8% | 23.5% (14.1 - 37.7) | 3.7% | 18.4% (11.5 - 28.7) |  |
| T4 | 1.5% | 21.9% (15.8 - 30.0) | 0.0% | 18.7% (11.3 - 30.1) | 3.2% | 25.4% (16.4 - 38.1) |  | 4.0% | 27.5% (17.3 - 41.9) | 0.0% | 18.4% (11.5 - 28.7) |  |
| T5 | 0.8% | 22.7% (16.5 - 30.9) | 0.0% | 18.7% (11.3 - 30.1) | 1.6% | 27.0% (17.7 - 39.8) |  | 1.9% | 29.4% (18.9 - 44.0) | 0.0% | 18.4% (11.5 - 28.7) |  |
| T6 | 3.3% | 26.0% (19.3 - 34.5) | 3.4% | 22.1% (13.9 - 34.1) | 3.2% | 30.2% (20.5 - 43.2) | 0.309 | 2.0% | 31.4% (20.5 - 46.0) | 4.3% | 22.7% (14.9 - 33.7) | 0.285 |
| **SAM by MUAC** |  |  |  |  |  |  |  |  |  |  |  |  |
|  | (n=160) |  | Bay (IDP population) n=71 |  | Hiran (Non-IDP population) n=63 |  |  | Rural (n=73) |  | Urban (n=87) |  |  |
| T1 | 8.8% | 8.8% (5.3 - 14.3) | 14.9% | 14.9% (8.5 - 25.2) | 3.5% | 3.5% (1.1 - 10.4) |  | 2.7% | 2.7% (0.7 - 10.5) | 13.8% | 13.8% (8.1 - 23.0) |  |
| T2 | 2.5% | 11.3% (7.3 - 17.3) | 4.1% | 19.0% (11.7 - 29.9) | 1.2% | 4.7% (1.8 - 11.9) |  | 0.0% | 2.7% (0.7 - 10.5) | 4.7% | 18.5% (11.7 - 28.3) |  |
| T3 | 1.2% | 12.5% (8.3 - 18.8) | 2.8% | 21.8% (14.0 - 33.1) | 0.0% | 4.7% (1.8 - 11.9) |  | 0.0% | 2.7% (0.7 - 10.5) | 2.3% | 20.8% (13.7 - 31.0) |  |
| T4 | 0.7% | 13.2% (8.8 - 19.5) | 0.0% | 21.8% (14.0 - 33.1) | 0.0% | 4.7% (1.8 - 11.9) |  | 0.0% | 2.7% (0.7 - 10.5) | 0.0% | 20.8% (13.7 - 31.0) |  |
| T5 | 0.0% | 13.2% (8.8 - 19.5) | 0.0% | 21.8% (14.0 - 33.1) | 1.1% | 5.8% (2.5 - 13.4) |  | 1.4% | 4.1% (1.3 - 12.2) | 0.0% | 20.8% (13.7 - 31.0) |  |
| T6 | 0.0% | 13.2% (8.8 - 19.5) | 0.0% | 21.8% (14.0 - 33.1) | 0.0% | 5.8% (2.5 - 13.4) | 0.003 * | 0.0% | 4.1% (1.3 - 12.2) | 0.0% | 20.8% (13.7 - 31.0) | 0.002 * |
| **Both SAM and MAM by WHZ** |  |  |  |  |  |  |  |  |  |  |  |  |
|  | (n=108) |  | Bay (IDP population) n=71 |  | Hiran (Non-IDP population) n=63 |  |  | Rural (n=39) |  | Urban (n=69) |  |  |
| T1 | 26.9% | 26.9% (19.5 - 36.3) | 27.4% | 27.4% (18.0 - 40.3) | 26.1% | 26.1% (15.7 - 41.3) |  | 28.2% | 28.2% (16.7 - 45.1) | 26.1% | 26.1% (17.3 - 38.2) |  |
| T2 | 9.3% | 36.2% (28.0 - 46.1) | 6.6% | 34.0% (23.7 - 47.3) | 13.0% | 39.1% (26.8 - 54.7) |  | 10.3% | 38.5% (25.3 - 55.5) | 8.9% | 35.0% (25.0 - 47.5) |  |
| T3 | 2.9% | 39.1% (30.6 - 49.0) | 3.4% | 37.4% (26.7 - 50.7) | 2.2% | 41.3% (28.7 - 56.8) |  | 2.5% | 41.0% (27.5 - 58.0) | 3.0% | 38.0% (27.7 - 50.6) |  |
| T4 | 4.9% | 44.0% (35.2 - 54.0) | 8.9% | 46.3% (34.7 - 59.7) | 0.0% | 41.3% (28.7 - 56.8) |  | 0.0% | 41.0% (27.5 - 58.0) | 7.9% | 45.9% (34.9 - 58.6) |  |
| T5 | 3.9% | 47.9% (38.9 - 57.9) | 1.8% | 48.1% (36.4 - 61.5) | 6.5% | 47.8% (34.6 - 63.0) |  | 7.7% | 48.7% (34.5 - 65.2) | 1.6% | 47.5% (36.4 - 60.1) |  |
| T6 | 2.2% | 50.1% (41.0 - | 2.2% | 50.3% (38.3 - | 2.2% | 50.0% (36.7 - | 0.991 | 2.6% | 51.3% (36.8 - | 1.9% | 49.4% (38.1 - | 0.860 |
|  |  | 60.0) |  | 63.7) |  | 65.1) |  |  | 67.5) |  | 62.0) |  |

For SAM relapse using MUAC, the cumulative incidence was notably lower, 13.2% (95% CI 8.8-19.5%) at T6. By month, incidence was highest within 1-3 timepoints at 8.8%, 2.5% and 1.2% respectively. Stratified analyses of wasting by MUAC are shown in **Table S2** but these should be interpreted with caution given small sample sizes and that MUAC severely underestimates the true burden of wasting and relapse.[29]

For combined SAM and MAM relapse by WHZ, the cumulative incidence was 50.1% (95% CI 41.0-60.0%) at T6, with substantial early relapse rates. Relapse was highest at T1 and T2 at 26.9% and 9.3% respectively. This then tapered to 2.9%, 4.9%, 3.9% and 2.2% from T3 to T6 respectively. Combined SAM and MAM relapse showed minimal variation across rural/urban settings (p=0.860) and regions (p=0.991). For combined SAM and MAM, there was no statistically significant difference by severity of WHZ at admission (p=0.335) or by age group (p=0.812).

### Relapse risk factors by WHZ

We investigated a range of potential predictors of SAM relapse using Cohort 1 (SAM relapse by WHZ), including admission characteristics, maternal factors, and child-specific variables **(Table 3)**. Both unadjusted and adjusted analyses were performed, with the adjusted model controlling for child age, sex, livelihood zone, and time to relapse.

**Table 3:**
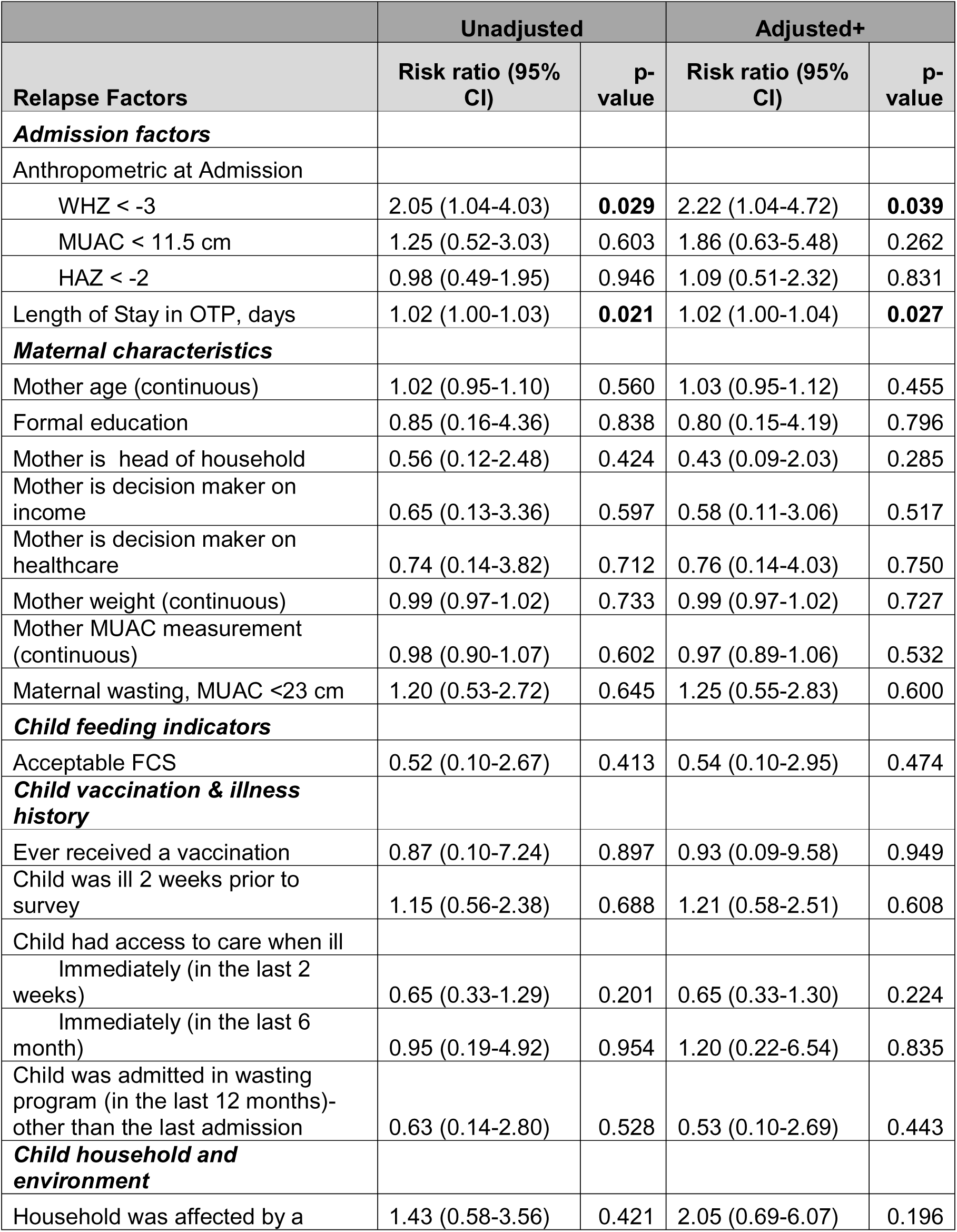
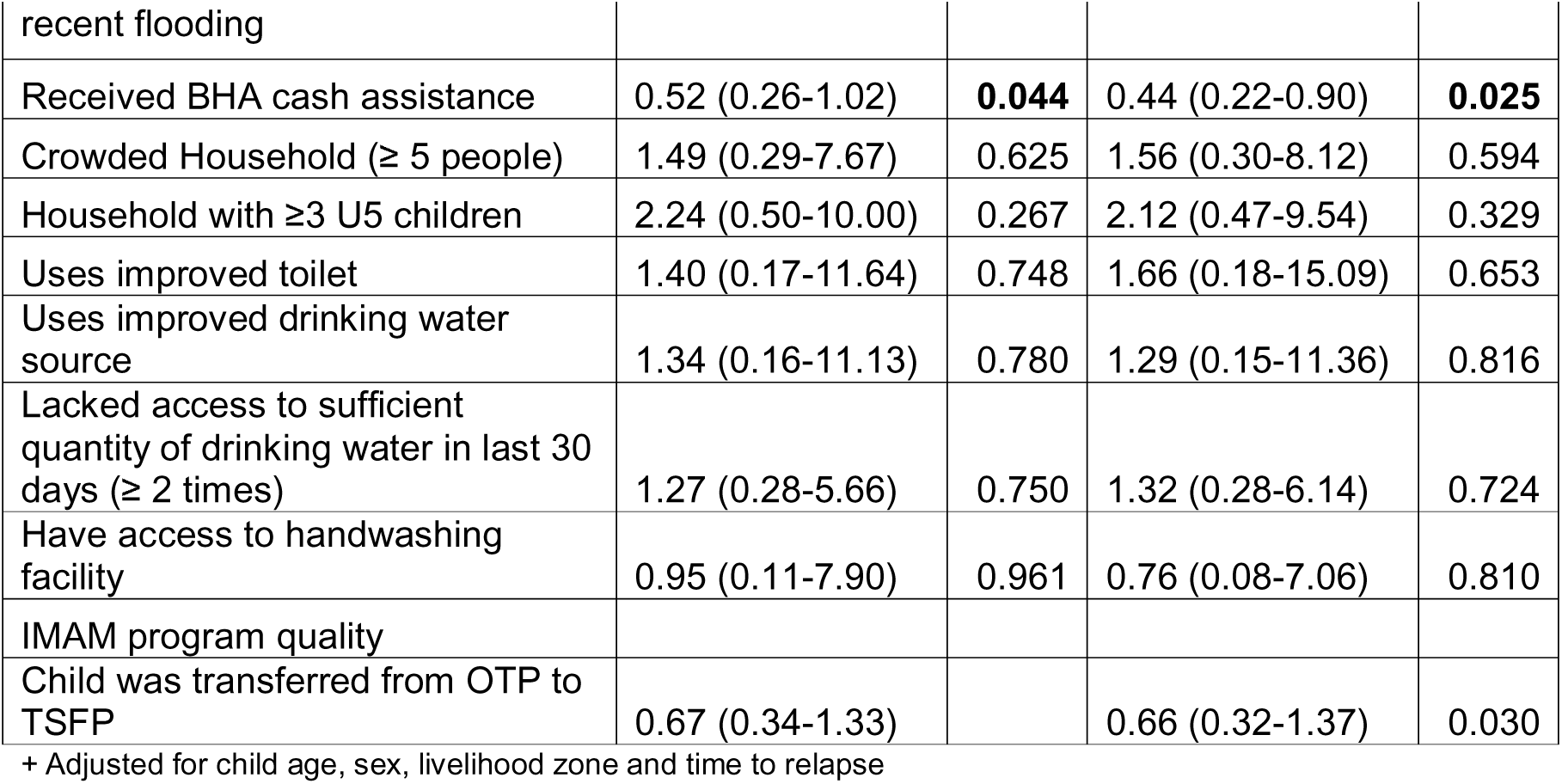
Risk ratios of individual and household factors associated with relapse to SAM after discharge from OTP treatment.

| Relapse Factors | Unadjusted |  | Adjusted+ |  |
| --- | --- | --- | --- | --- |
|  | Risk ratio (95% CI) | p-value | Risk ratio (95% CI) | p-value |
| **Admission factors** |  |  |  |  |
| Anthropometric at Admission |  |  |  |  |
| WHZ < -3 | 2.05 (1.04-4.03) | **0.029** | 2.22 (1.04-4.72) | **0.039** |
| MUAC < 11.5 cm | 1.25 (0.52-3.03) | 0.603 | 1.86 (0.63-5.48) | 0.262 |
| HAZ < -2 | 0.98 (0.49-1.95) | 0.946 | 1.09 (0.51-2.32) | 0.831 |
| Length of Stay in OTP, days | 1.02 (1.00-1.03) | **0.021** | 1.02 (1.00-1.04) | **0.027** |
| **Maternal characteristics** |  |  |  |  |
| Mother age (continuous) | 1.02 (0.95-1.10) | 0.560 | 1.03 (0.95-1.12) | 0.455 |
| Formal education | 0.85 (0.16-4.36) | 0.838 | 0.80 (0.15-4.19) | 0.796 |
| Mother is head of household | 0.56 (0.12-2.48) | 0.424 | 0.43 (0.09-2.03) | 0.285 |
| Mother is decision maker on income | 0.65 (0.13-3.36) | 0.597 | 0.58 (0.11-3.06) | 0.517 |
| Mother is decision maker on healthcare | 0.74 (0.14-3.82) | 0.712 | 0.76 (0.14-4.03) | 0.750 |
| Mother weight (continuous) | 0.99 (0.97-1.02) | 0.733 | 0.99 (0.97-1.02) | 0.727 |
| Mother MUAC measurement (continuous) | 0.98 (0.90-1.07) | 0.602 | 0.97 (0.89-1.06) | 0.532 |
| Maternal wasting, MUAC <23 cm | 1.20 (0.53-2.72) | 0.645 | 1.25 (0.55-2.83) | 0.600 |
| **Child feeding indicators** |  |  |  |  |
| Acceptable FCS | 0.52 (0.10-2.67) | 0.413 | 0.54 (0.10-2.95) | 0.474 |
| **Child vaccination & illness history** |  |  |  |  |
| Ever received a vaccination | 0.87 (0.10-7.24) | 0.897 | 0.93 (0.09-9.58) | 0.949 |
| Child was ill 2 weeks prior to survey | 1.15 (0.56-2.38) | 0.688 | 1.21 (0.58-2.51) | 0.608 |
| Child had access to care when ill |  |  |  |  |
| Immediately (in the last 2 weeks) | 0.65 (0.33-1.29) | 0.201 | 0.65 (0.33-1.30) | 0.224 |
| Immediately (in the last 6 months) | 0.95 (0.19-4.92) | 0.954 | 1.20 (0.22-6.54) | 0.835 |
| Child was admitted in wasting program (in the last 12 months)- other than the last admission | 0.63 (0.14-2.80) | 0.528 | 0.53 (0.10-2.69) | 0.443 |
| **Child household and environment** |  |  |  |  |
| Household was affected by a | 1.43 (0.58-3.56) | 0.421 | 2.05 (0.69-6.07) | 0.196 |
| recent flooding |  |  |  |  |
| Received BHA cash assistance | 0.52 (0.26-1.02) | **0.044** | 0.44 (0.22-0.90) | **0.025** |
| Crowded Household ($\geq 5$ people) | 1.49 (0.29-7.67) | 0.625 | 1.56 (0.30-8.12) | 0.594 |
| Household with $\geq 3$ U5 children | 2.24 (0.50-10.00) | 0.267 | 2.12 (0.47-9.54) | 0.329 |
| Uses improved toilet | 1.40 (0.17-11.64) | 0.748 | 1.66 (0.18-15.09) | 0.653 |
| Uses improved drinking water source | 1.34 (0.16-11.13) | 0.780 | 1.29 (0.15-11.36) | 0.816 |
| Lacked access to sufficient quantity of drinking water in last 30 days ($\geq 2$ times) | 1.27 (0.28-5.66) | 0.750 | 1.32 (0.28-6.14) | 0.724 |
| Have access to handwashing facility | 0.95 (0.11-7.90) | 0.961 | 0.76 (0.08-7.06) | 0.810 |
| IMAM program quality |  |  |  |  |
| Child was transferred from OTP to TSFP | 0.67 (0.34-1.33) |  | 0.66 (0.32-1.37) | 0.030 |

+ Adjusted for child age, sex, livelihood zone and time to relapse

In the adjusted analysis, three factors emerged as significant predictors of SAM relapse. Children with a WHZ below −3 SD at admission had a statistically significantly higher risk of relapse (adjusted HR: 2.22, 95% CI: 1.04-4.72, p=0.039). Similarly, a longer duration of OTP stay was associated with an increased relapse risk (adjusted HR: 1.02 per day, 95% CI: 1.00-1.04, p=0.027). Additionally, participation in the BHA-funded cash program was protective against relapse (adjusted HR: 0.44, 95% CI: 0.22-0.90, p=0.025). Other factors examined, including maternal characteristics (e.g., age, education, decision-making power), child feeding practices, vaccination history, and household environment did not show statistically significant associations with relapse risk in the adjusted analysis.

## DISCUSSION

To our knowledge, this is the first study of child wasting relapse rates and determinants in the Bay and Hiran regions of Somalia. We found that incidence of SAM at T6 post-OTP exit was as high as 26.0% (22.1% in Bay and 30.2% in Hiran), and incidence of MAM was 50.1% (50.3% in Bay and 50.0% in Hiran). These rates are higher compared to those found in a recent multi-country study, where a 5% cumulative incidence of SAM or all-cause death and 23% acute malnutrition or all-cause death was reported among post-SAM CU5 in Mogadishu, Somalia at 6 months when employing a MUAC-only acute malnutrition criterion [9]. It’s important to note that data for our study was collected between November to April which overlap the raining season (deyr) and dry season (Jilaal), while in the multi-country study, data collection occurred during “Gu”, the main raining season between harvest (April-June). In our study, the incidence of SAM was highest in the first 3 follow-up time points and decreased afterwards; similar findings have been reported elsewhere, where the majority of relapse cases occurred in the first 3 months of follow-up [30]. On the other hand, in a multi-country study that included Somalia, the rate of relapse remained relatively constant over the six-month period at a relapse incidence rate of 1 case per 100 child-months [9]. Our results of higher incidence of relapse initially during follow-up could be due to multiple contextual factors, including seasonality and major flooding that occurred early on in the follow-up period and affected livelihoods and food security in Bay and Hiran [31]. Children in our cohort continued to relapse to wasting throughout the study period, indicating that relapse risk in Somalia persists over time and children may be at risk both immediately following discharge and for at least 6 months afterwards, which aligns with previous literature in Somalia and globally [9]. This finding has policy and programmatic implications for ensuring that children’s nutrition, health, and food security are protected and promoted not only through initial wasting recovery, but also through longer-term investments to prevent relapse.

The findings from this analysis demonstrate that relapse rates in the Bay and Hiran regions of Somalia vary by anthropometric indicator, by region, and by demographic factors. When defined by WHZ, the incidence of SAM relapse was 26% and relapse rate was higher in rural areas (31.4% in rural areas vs 22.7% in urban areas), but not statistically significant (p=0.285). MUAC relapse rate is lower when compared to WHZ at all follow-up timepoints. Variable relapse rates according to anthropometric criteria have also been reported elsewhere, with one study finding that using a MUAC < 115 mm or nutritional edema criteria missed over 80% of relapse cases in their sample [9,14],[29]. These differences in wasting and relapse burden according to measurement metric underscore the critical importance of using a reliable and accurate measure of wasting, such as WHZ, where possible; MUAC alone may underestimate and misclassify wasting risk – an issue that cannot be understated [29].

In our analysis, WHZ at admission to OTP was statistically significantly associated with risk of relapse among children under 5. This finding is consistent with previous results that have also shown anthropometry at admission to be associated with acute malnutrition relapse among CU5 [6,9,13,16]. We also found length of stay in the OTP to be associated with wasting relapse risk in our cohort. This finding is consistent with work done in Mali, where researchers likewise found a longer length of stay to be predictive of relapse, and one hypothesis for this association with relapse outcomes is that children staying longer in OTP may have other complications and poorer health in general, resulting in increased nutritional vulnerability even after OTP discharge [13]. In our analysis, participation in the BHA cash assistance program was protective against relapse. This finding is consistent with previous work from the Democratic Republic of the Congo where receiving cash transfers was associated with decreased rates of relapse [6,32]. Interestingly, other factors such as maternal characteristics (e.g., age, education, and decision-making power), child feeding practices, vaccination history, and household environment did not show statistically significant associations with relapse risk in our adjusted analysis. However, additional work done by our group has identified factors such as maternal age, MUAC, and education, household WASH environment, and child vaccination and diet as drivers of wasting in the Bay and Hiran regions [33]. Additionally, previous work in Somalia has shown that living in a household experiencing severe hunger or falling into the lowest wealth quartile were risk factors for relapse in post-SAM children [9]. Other analyses in sub-Saharan Africa have found such factors as child sickness and vaccination, child dietary diversity, maternal/caretaker age, maternal literacy, hygiene practices, and household water source and food insecurity to be associated with acute malnutrition relapse among CU5 [6,13,15,16,24,34,35]. In light of contrasting evidence, differing definitions of relapse, and the known relationship between factors such as food security and nutrition outcomes, our findings on determinants of relapse risk should be interpreted cautiously given the relatively wide confidence intervals for some estimates.

Our findings highlight critical needs across the continuum of SAM care given 49% of children who exited OTP were not transferred to TSFP in our study. Treatment services require strengthening, with evidence from Ethiopia showing poor adherence to guidelines and inadequate staffing [36]. Discharge practices need improvement, as demonstrated by premature discharge cases in our study and others [6],[12], [13], [15],[9], [34]. Post-discharge monitoring must be enhanced through community-based approaches and early warning systems. While family-MUAC monitoring was implemented in Somalia in 2020 [37], additional approaches like weekly MUAC reporting and regular follow-up are needed [6]. For prevention, while SQ-LNS shows promise for preventing child wasting, research on its utility for prevention of wasting relapse is limited [38], [39].

The disparity in relapse rates by anthropometric criteria (26% WHZ vs 13.2% MUAC) underscore the need for standardized definitions and measurements of wasting relapse [6], [29],[40]. This standardization is crucial for effective resource allocation and treatment planning in humanitarian settings.

Our study was limited by smaller-than-planned recruitment due to access constraints, though the sample size (>30) was adequate for our analyses [41]. While lacking a non-post-SAM control group, our diverse sample allowed assessment of relapse patterns across different subpopulations. We propose future studies that extend beyond 7 months of follow-up, as evidence shows children remain at risk of poor outcomes over longer periods [6], [42]. Larger, longer-term studies across multiple regions of Somalia would enable analysis of seasonal trends in relapse [43] and strengthen understanding of relapse determinants.

Research priorities include evaluating relapse prevention approaches like SQ-LNS, Blanket Supplementary Feeding Program (BSFP), Cash and discharge criteria modification [6] [38], and developing standardized definitions of relapse to improve measurement consistency and resource utilization [40], [29]. Well-powered studies across diverse populations are needed to better characterize both relapse rates and their underlying causes throughout Somalia.

## CONCLUSIONS

With SAM relapse rates as high as 26% and MAM as high as 50% at seven months among children, it is without a doubt clear that wasting relapse is a serious public health concern in Somalia. Moreover, the burden that relapse places on humanitarian resources by repeatedly treating the same children and the risk that relapse poses to children’s health and survival, suggests that addressing relapse should be top of funding, policy and programming priorities in the country. The key drivers of relapse identified in this study shed some light on ways to intervene however further research is needed. Additionally, the variable rates of relapse according to anthropometric criteria emphasizes the need for future work to adopt an accurate and reliable definition of wasting relapse for more effective resource distribution and identification and treatment of relapsed children.

## Ethics statements

Informed consent was obtained from all study participants. The study was conducted in accordance with the Declaration of Helsinki and approved by the Institutional Review Board of the Johns Hopkins Bloomberg School of Public Health (protocol code 24476, date of approval: May 17, 2023), the Ministry of Health and Human Services in Somalia (XAG/203/23, date of approval: April 26th, 2023), and the Save the Children International Ethics Review Committee.

## Data Availability

This data was collected on a vulnerable population linked to a large scale cash assistance program in Somalia. The data may be made available upon request with permission from local study investigators.

## Authorship contributions

Conceptualization (K.K.A., S.W.,S.G., S.G., N.A.,S.A.M., M.A.N., M.O.,A.A.,Q.K.,L.S.,M.T.,F.L., A.S.,M.B.,M.O.,P.S.,F.M.,A.A.M.,A.A.A.,A.A.), data collection (K.K.A., S.W.,S.G., S.G., N.A.,S.A.,M.B.,A.A.); data cleaning and analysis (K.K.A., S.W.,S.G., S.G., N.A.); draft manuscript preparation (K.K.A., S.W.,S.G., S.G., N.A.); critical review and editing of manuscript (K.K.A., S.W.,S.G., S.G., N.A.,S.A.M., M.A.N., M.O.,A.A.,Q.K.,L.S.,M.T.,F.L., A.S.,M.B.,M.O.,P.S.,F.M.,A.A.M.,A.A.A.,A.A.); final approval of manuscript (K.K.A., S.W.,S.G., S.G., N.A., S.A.M., M.A.N., M.O., A.A., Q.K., L.S., M.T., F.L., A.S., M.B., M.O.,P.S.,F.M.,A.A.M.,A.A.A.,A.A.). All authors have read and agree to the published version of this manuscript and agree to be accountable for all aspects of the work.

## Acknowledgements

We are grateful for the support of Bethan Mathias and Emmanuel Falade, and all Save the Children Somalia that participated in the study. Without their input and the support of the Somalia Ministry of Health and Nutrition, this work would not have been possible.

