## Supplemental Table S1 and S2 for "Levels and determinants of child wasting relapse: a prospective cohort study from Somalia"

### S1: Relapse Study Questionnaire

| |
|---|
| **Module A: Enumerator and location details** |
| Enumerator Team |
| Household Unique ID |
| Mother's Name |
| Mother's Age |
| Index Child Unique ID |
| Is the Index Child a singleton birth? (i.e. not a twin or triplet) |
| Date Index Child was Discharged from OTP |
| Displacement Status |
| Select region from list. |
| Select district from list. |
| Select village from list. |
| **Informed Consent** |
| Thank you for the opportunity to speak with you. We are from Save the Children. We are conducting a survey with randomly selected program households to learn more about who and why children who are discharged from OTP treatment re-develop malnutirion. The questions will take about <span style="color:red">45 minutes</span> to complete. Your participation is voluntary. If you agree to participate, you can choose to stop at any time or skip any questions you do not want to answer. There will be no compensation for participating in this survey. If you do not agree to participate, it will not affect your status in the program or the services you receive. Any personal information such as names that you share will be kept confidential and only a limited number of Save the Children staff will have access to it. We intend to share the resulting analysis, which will not include personal information, with our donors and partners to help improve future programs. Let me know if you have any questions about this. |
| What is your relationship to the index child in this household? |
| Specify relationship to the index child. |
| Is your household in the SCI BHA program? |
| **Section 1: Respondent information** |
| What is the highest level of school you attended, including madrasa? |
| Are you the head of the household? |
| If no, what is the gender of the head of household? |
| What is your relationship with the head of the household? |
| Who usually decides how the household income/earnings (all sources of income) will be used: you, your husband, or you and your husband jointly? |
| If other, please specify. |
| Who usually makes decisions about healthcare for yourself: you, your husband, or you and your husband jointly? |
| If other, please specify. |
| Is any member of your household <span style="font-weight:bold">currently receiving any assistance from any institution</span> such as the government, international organizations, religious bodies in the form of...? |
| If other, please specify. |
| How much did you receive (either total value in USD or kg or other unit)? |
| How oftern did you receive it? (weekly, monthly, etc) |
| **Basic Causes: Context, Capital, Resources** |
| Does your household own any of the following livestock? |
| How many camels does your household currently own? |
| How many cattle does your household currently own? |
| How many goats does your household currently own? |
| How many donkeys does your household currently own? |
| How many horses does your household currently own? |
| How many poultry does your household currently own? |
| Does any member of this household own any agricultural land? |
| How many hectares of agricultural land do members of this household own? |
| Does your household have any of the folowing? |

| |
|---|
| Does any member of this household own: |
| Does any member of this household have a bank account? |
| Does any member of this household use a mobile phone to make financial transactions such as sending or receiving money, paying bills, purchasing goods or services, or receiving wages? |
| How many rooms in this household are used for sleeping? |
| What is the main material of the floor of the house? |
| If other, please specify. |
| What is the main material of the roof of the house? |
| If other, please specify. |
| What is the main material of the exterior walls of the house? |
| If other, please specify. |
| Does your household have any mosquito bednets? |
| In your household, how do you get the money to cover your household needs/expenses? |
| **Household Composition, Morbidity and Migrations** |
| **Child Anthropometrics** |
| How many of the children in this household are between 0 to 59 months ? |
| Index Child U5 |
| <5yr kids for Mother/Carer |
| Index Child Name |
| What is the sex of the index child ${child}? |
| What is the index child ${child}'s age in months? |
| How did you confirm the age of the index child ${child}? |
| Was the index child enrolled in a wasting treatment program in the last 12 months other than the most recent time? |
| If yes, was it OTP or TSFP? |
| About how many weeks were they enrolled? |
| Were they discharged from the wasting treatment program because they recovered? |
| Were any other under-five children in the household enrolled in a treatment program in the last 12 months? |
| Has the index child ${child} received a deworming tablet within the last 6 months? |
| ${child} Index Child Weight (kg) to the nearest 0.1kg |
| ${child} Index Child Height (cm) to the nearest 0.1cm |
| ${child} Oedema present in the index child |
| ${child} Index Child MUAC (cm) to the nearest 0.1 |
| Lt5 position |
| Now I am going to take your MUAC measurement from your arm. |
| Weight (kg) to the nearest 0.1kg |
| Height (cm) to the nearest 0.1cm |
| How many rooms in this household are used for sleeping? |
| How many people slept in household last night? |
| Record the type of dwelling the index child's household is living in |
| **Underlying causes - Inadequate care and feeding practices** |
| **Child Vaccination History** |
| Thinking about the index child, ${child}, did s/he ever receive any vaccinations to prevent diseases, including vaccinations received in campaigns or immunization days or child health days? |
| Has the index child ${child} ever received any of the following vaccinations? |
| How many times did the index child ${child} receive the pentavalent vaccine? |
| How many times did the index child ${child} receive the measles vaccine? |
| **Child Health & Nutrition** |
| Has the index child ${child} had diarrhea, vomiting, fever,or cough in the last 2 weeks? |
| Has the index child ${child} had malaria or TB in the last 2 weeks? |
| Has the index child ${child} lost their appetite in the last 2 weeks? |
| Did you seek treatment for the illness from any source? |
| Where did you seek treatment from? |
| If other, please specify. |

| |
|---|
| In the last 2 weeks, if the index child needed health care because of any symptom of illness, did you seek care immediately or was there a delay or did you not seek care? |
| What are some reasons you delayed or didn't seek care? |
| If other, please specify. |
| Now thinking about the last 6 months, when the index child needed health care because of any symptom of illness (apart from the wasting treatment), did they receive care or was it delayed? |
| What are some reasons you delayed or didn't seek care? |
| If other, please specify. |
| Does index child have any chronic medical condition? |
| If yes, record chronic medical condition |
| Does index child have any physical disability? |
| If yes, record nature of physical disability |
| **Minimum Dietary Diversity for Children (MDD-C)** |
| Now I'm going to ask you some questions about the index child's eating and drinking during the last 24 hours (day and night). |
| Has the index child ${child} ever been breastfed? |
| How long after birth did you put the index child ${child} to the breast for the first time? |
| Note: If the answer is less than 1 hour, write '00' for the hours. If it is 1-24 hours, record the number of hours. |
| Record the number of days. |
| Was the index child ${child} breastfed yesterday during the day or at night? This includes any breast milk the child drank from a bottle, cup, or spoon as well as directly from the breast. |
| If yes, how many times did you breastfeed the index child? |
| Now I would like to ask you about some medicines and vitamins that are sometimes given to infants.<br>Was the index child ${child} given any vitamin drops or other medicines as drops yesterday during the day or at night? |
| Was the index child ${child} given oral rehydration solution yesterday during the day or at night? |
| Next I would like to ask you about some liquids that the index child ${child} may have had yesterday during the day or at night. Did ${child} have any of the following... (read list) |
| Plain water? |
| Infant formula? |
| How many times yesterday during the day or at night did the index child ${child} consume any formula? |
| Any milk such as tinned, powdered, or fresh animal milk? |
| How many times yesterday during the day or at night did the index child ${child} consume any of these types of milk? |
| Any juice or juice drinks? |
| Clear broth? |
| Yogurt? |
| How many times yesterday during the day or at night did the index child ${child} consume any yogurt? |
| Any thin porridge? |
| Any other liquids? |
| Did the index child ${child} drink anything from a bottle with a nipple yesterday during the day or night? |
| Do you remember how old the index child ${child} was when you started feeding them liquids for the first time? |
| Can you tell me how old the index child ${child} was in months? |
| At what age should you start to feed your child liquids for the first time? (in months) |
| Please describe everything that the index child ${child} ate yesterday during the day or night, whether at home or outside the home. Did s/he have any... (read from list) |
| Food made from grains, such as bread, rice, noodles, porridge |
| Pumpkin, carrots, squash, or sweet potatoes that are yellow or orange inside |
| White potatoes, white yams, manioc, cassava, or any other foods made from roots |
| Any dark green leafy vegetables |
| Any other vegetables? |
| Ripe mangoes, ripe papayas, or other local vitamin A-rich fruits |
| Any other fruits? |
| Liver, kidney, heart, or other organs from domesticated animals such as cow, goat, chicken or duck |
| Any meat from domesticated animals such as beef, camel, lamb, goat, chicken, or duck |

| |
|---|
| Liver, kidney, heart, or other organs from wild animals, such as birds, wild pigeons, wild fowl, wild boar, rodents, wild goat |
| Any flesh from wild animals, such as birds, wild pigeons, wild fowl, |
| Eggs |
| Fresh or dried fish, shellfish, or seafood |
| Any foods made from beans, peas, lentils, peanuts, peanut paste or other legumes |
| Any foods made from nuts and seeds such as pumpkin seeds, cashews, jackfruit |
| Cheese, yogurt, or other milk products |
| Any oil, fats, or butter, or foods made with any of these |
| Any sugary foods such as chocolates, sweets, candies, pastries, cakes, or biscuits |
| Condiments for flavor, such as chilies, spices, herbs, or fish powder |
| crubs, snails, |
| Foods made with red palm oil, red palm nut, or red palm nut pulp sauce |
| Did the index child ${child} eat any solid, semi-solid, or soft foods yesterday during the day or at night? |
| If yes, how many times? |
| Do you remember how old the index child ${child} was when you started feeding them solid food for the first time? |
| Can you tell me how old the index child ${child} was in months? |
| At what age should you start to feed your child solids for the first time? (in months) |
| **Wasting Treatment Program Experience** |
| Now I'm going to ask you questions about the treatment the index child received for wasting (Emphasize that these questions are for the latest treatment the child received in cases where child had many wasting treatments previously) This will only take a couple of minutes. |
| Can you please tell me why your child was enrolled in a treatment program? (open-ended) |
| When was index child enrolled in the OTP? |
| How many times did you go to OTP |
| Were you discharged/transferred from OTP to TSFP? |
| If yes, When were you transferred to TSFP? |
| How many times did you go to TSFP after being transferred? |
| When were you discharged from TSFP? |
| How long does it take you to get to the treatment center by walking? (one way walking distance) |
| How do you usually get to the treatment center? Please tell me your primary mode of transportation. |
| During the treatment program, what services did you/the index child receive? |
| If other, please specify. |
| How many times did you/the index child receive RUTF? |
| How many times did you/the index child receive RUSF? |
| How many times did you/the index child receive CSB++? |
| How many times did you/the index child receive Amoxicillin? |
| How many times did you/the index child receive Albendazole/Mebendazole? |
| How many times did you/the index child receive Measles Vaccine? |
| How many times did you/the index child receive Vitamin A supplements? |
| How many times did you receive Training on the causes of wasting? |
| Were you satisfied with the service/treatment received? |
| If your friend or neighbor's child was ill, would you recommend this service? |
| If no, why? (open-ended) |
| **Food Consumption Score (FCS)** |
| Now I would like to ask you about all the different foods that your WHOLE household have eaten in the last 7 days. This will only take two minutes and we are almost finished.<br>Could you please tell me how many days in the past week you and your household has eaten the following foods? |
| cereals_tubers |
| In the past week, approximately how many days has your household eaten cereals and tubers such as maize, maize porridge, rice, sorghum, millet, pasta, bread and other cereals, cassava/yucca, potato, or sweet potato? |
| legumes_group |
| In the past week, approximately how many days has your household eaten legumes and nuts such as beans, peas, groundnuts or cashew nuts? |
| vegetables_group |

| |
|---|
| In the past week, approximately how many days has your household eaten vegetables or leaves? |
| fruits_group |
| In the past week, approximately how many days has your household eaten fresh fruits? |
| meat_fish_group |
| In the past week, approximately how many days has your household eaten eggs or meat such as beef, goat, poultry, camel, or fish? |
| dairy_group |
| In the past week, approximately how many days has your household eaten dairy products such as milk, yogurt, or cheese? |
| sugar_group |
| In the past week, approximately how many days has your household eaten sugar and sugary products such as honey, jam, candy, or pastries. |
| oil_group |
| In the past week, approximately how many days has your household eaten oils, fats or butter? |
| condiment_group |
| In the past week, approximately how many days has your household eaten condiments such as spices, tea, coffee, salt, fish powder, or small amounts of milk for tea? |
| **Section 5: Underlying causes - Unhealthy HH environment** |
| Lastly, I would like to ask you just a few questions about your household's water needs and consumption, and hygiene habits. This will take about two minutes and we are done after this set of questions. |
| What is the main source of drinking water for members of your household? |
| If other, specify. |
| In the last 30 days, how frequently have you or your household <span style="font-weight:bold">not had sufficient</span> drinking water? |
| What was the reason that you were unable to access water in sufficient quantities when needed? (select all that apply) |
| If other, please specify. |
| What is usually done to make the water safer to drink? (select all that apply) |
| If other, please specify. |
| Does the household have access to a handwashing place that has soap and water? |
| What type of handwashing station is it? |
| If other, specify |
| Is water present for handwashing? |
| Is cleansing agent available for handwashing? |
| What kind of toilet facility do members of your household usually use? |
| If other, specify. |

**S2:** Incidence of Wasting for SAM (By WHZ), SAM (By MUAC) and MAM (By WHZ) at each follow-up point disaggregated by child characteristics

| | Age | | | | | Severity of Wasting at Admission | | | | |
|---|---|---|---|---|---|---|---|---|---|---|
| Follow-up Time | Incidence | Cumulative Incidence (95% CI) | Incidence | Cumulative Incidence (95% CI) | p-value | Incidence | Cumulative Incidence (95% CI) | Incidence | Cumulative Incidence (95% CI) | p-value |
| SAM by WHZ | | | | | | | | | | |
| | *< 2 years (n=76)* | | *≥ 2 years (n=58)* | | | *WHZ <-3SD (n=40)* | | *WHZ ≥ -3SD (n=94)* | | |
| T1 | 5.3% | 5.3% (2.0 - 13.4) | 5.2% | 5.2% (1.7 - 15.2) | | 12.5% | 12.5% (5.4 - 27.5) | 2.1% | 2.1% (0.5 - 8.2) | |
| T2 | 10.8% | 16.1% (9.5 - 26.6) | 6.9% | 12.1% (5.9 - 23.7) | | 15.0% | 27.5% (16.3 - 44.1) | 6.6% | 8.7% (4.4 - 16.6) | |
| T3 | 2.7% | 18.8% (11.6 - 29.7) | 10.3% | 22.4% (13.7 - 35.5) | | 4.8% | 32.3% (20.2 - 49.1) | 6.6% | 15.3% (9.3 - 24.4) | |
| T4 | 1.4% | 20.2% (12.7 - 31.3) | 1.7% | 24.1% (15.1 - 37.3) | | 2.5% | 34.8% (22.3 - 51.5) | 1.1% | 16.4% (10.2 - 25.7) | |
| T5 | 0.0% | 20.2% (12.7 - 31.3) | 1.8% | 25.9% (16.5 - 39.2) | | 0.0% | 34.8% (22.3 - 51.5) | 1.1% | 17.5% (11.1 - 27.0) | |
| T6 | 3.1% | 23.3% (15.1 - 34.8) | 3.6% | 29.5% (19.5 - 43.1) | 0.475 | 2.6% | 37.4% (24.5 - 54.1) | 3.7% | 21.2% (14.0 - 31.2) | 0.029* |
| SAM by MUAC | | | | | | | | | | |
| | *< 2 years (n=82)* | | *≥ 2 years (n=78)* | | | *MUAC ≤11.2cm (n=72)* | | *MUAC >11.2cm (n=88)* | | |
| T1 | 11.0% | 11.0% (5.9 - 20.0) | 6.4% | 6.4% (2.7 - 14.7) | | 11.1% | 11.1% (5.7 - 21.0) | 6.8% | 6.8% (3.1 - 14.6) | |
| T2 | 4.9% | 15.9% (9.6 - 25.8) | 0.0% | 6.4% (2.7 - 14.7) | | 2.8% | 13.9% (7.7 - 24.3) | 2.3% | 9.1% (4.7 - 17.4) | |
| T3 | 1.3% | 17.2% (10.5 - 27.3) | 1.3% | 7.7% (3.5 - 16.4) | | 1.4% | 15.3% (8.8 - 25.9) | 1.2% | 10.3% (5.5 - 18.8) | |
| T4 | 0.0% | 17.2% (10.5 - 27.3) | 0.0% | 7.7% (3.5 - 16.4) | | 0.0% | 15.3% (8.8 - 25.9) | 0.0% | 10.3% (5.5 - 18.8) | |
| T5 | 1.2% | 18.4% (11.6 - 28.7) | 0.0% | 7.7% (3.5 - 16.4) | | 1.5% | 16.8% (9.9 - 27.6) | 0.0% | 10.3% (5.5 - 18.8) | |
| T6 | 0.0% | 18.4% (11.6 - 28.7) | 0.0% | 7.7% (3.5 - 16.4) | 0.048* | 0.0% | 16.8% (9.9 - 27.6) | 0.0% | 10.3% (5.5 - 18.8) | 0.233 |
| Both SAM and MAM by WHZ | | | | | | | | | | |
| | *< 2 years (n=65)* | | *≥ 2 years (n=43)* | | | *WHZ <-3SD (n=23)* | | *WHZ ≥ -3SD (n=85)* | | |
| T1 | 27.7% | 27.7% (18.4 - 40.3) | 25.6% | 25.6% (15.1 - 41.4) | | 26.1% | 26.1% (12.7 - 49.1) | 27.1% | 27.1% (18.9 - 37.9) | |
| T2 | 6.3% | 34.0% (23.9 - 46.9) | 13.9% | 39.5% (26.7 - 55.7) | | 4.3% | 30.4% (15.8 - 53.4) | 10.7% | 37.8% (28.5 - 49.1) | |
| T3 | 3.1% | 37.1% (26.6 - 50.1) | 2.4% | 41.9% (28.9 - 58.1) | | 4.7% | 35.1% (19.3 - 58.1) | 2.4% | 40.2% (30.7 - 51.5) | |
| T4 | 8.3% | 45.4% (34.1 - 58.4) | 0.0% | 41.9% (28.9 - 58.1) | | 0.0% | 35.1% (19.3 - 58.1) | 6.2% | 46.4% (36.5 - 57.7) | |
| T5 | 1.7% | 47.1% (35.7 - 60.0) | 7.3% | 49.2% (35.4 - 65.0) | | 0.0% | 35.1% (19.3 - 58.1) | 5.0% | 51.4% (41.2 - 62.5) | |
| T6 | 1.8% | 48.9% (37.3 - 61.8) | 2.7% | 51.9% (37.9 - 67.5) | 0.800 | 5.4% | 40.5% (23.3 - 63.7) | 1.4% | 52.8% (42.5 - 63.8) | 0.335 |
